# Inequalities in initiation of COVID19 vaccination by age and population group in Israel-December 2020-April 2021

**DOI:** 10.1101/2021.06.14.21258882

**Authors:** Yanay Gorelik, Michael Edelstein

## Abstract

**Background:** In Israel, COVID19 vaccination coverage varies widely by population group and age. Despite the vaccine being locally and freely available in the entire country. Comparing crude coverage between localities and population groups is misleading because of differing age structures in different population groups. In order to describe inequalities in COVID19 vaccine initiation we determined differences in age-specific dose 1 vaccine coverage between the main population groups in Israel, and characterised the influence of age on vaccine coverage within each of these groups.

**Methods:** We obtained daily doses administered by municipality and age from the Ministry of Health, and demographic data from the Central Bureau of Statistics. We determined whether the relative proportion of people vaccinated in each age group (15-19, 20-29, 30-39, 40-49, 50-59, 60+) changed by population group (General Jewish, Ultra-Orthodox and Arab) using ANOVA and quantified association between age, population group and vaccine coverage using binomial regression.

**Results:** 8,507,723 individuals in 268 localities were included. Compared with the General Jewish population, vaccine coverage was lower among the Arab and Ultra-Orthodox populations and lowest in the Ultra-Orthodox population in all age groups. Gaps between population groups differed according to age group (p<0.001). In all populations, coverage decreased with decreasing age (p<0.001). The Ultra-orthodox population was the least vaccinated in all age groups relatively to those aged 60 and over

**Conclusions:** In all age groups, the Ultra-Orthodox population had the lowest vaccine coverage. The younger the age group, the more Ultra-Orthodox Jews are diverging from their age peers in terms of initiating COVID19 vaccination. These findings suggest generational differences in terms of vaccination behaviour in this group. Qualitative studies understanding the causes behind this divergence can inform tailored vaccination strategies.

## Introduction

By April 2021, The COVID-19 pandemic, which was detected in China in December 2019 and rapidly spread globally, had infected over 136 million and killed at least 2.9 million individuals^1^, disrupting trade, travel, education, and the economy on an unprecedented scale. Despite a range of measures such as social distancing, masks, mass testing, surveillance and repeated lockdowns, implemented to varying degrees of stringency by almost every country in the world, the pandemic continued to spread. The emergence of safe and effective vaccines in December 2020 (^2, 3, 4^), with more at various stages of development^5^ offer new hope for controlling the pandemic.

Israel has not been spared by the pandemic and as of April 11 2021 reported 835,871 cases and 6,294 deaths^6^. By this date Israel had vaccinated 57.1% of the population with one dose of the BNT16b2 mRNA vaccine and 52.9% with the second dose, including over 92% of individuals over the age of 60^6^, the highest proportion in the world^6^. The benefits of such high vaccine coverage are being seen, with a sharp decrease in transmission and incidence since March 2020^6^ despite a gradual easing of control measures. Reasons for this success include Israel’s small size, a centralized national system of government, experience in managing large-scale national emergencies, the organizational, IT and logistical capacities of Israel’s community-based health care providers, and the mobilization of special government funding for vaccine purchase and distribution.^7^

Vaccinations against COVID in Israel take place in community health centres (HMOs), hospitals, ad-hoc vaccination centres in repurposed facilities such as stadiums and sport centres, and mobile units reaching remote and small communities. Those over the age of 60 first became eligible on 20/12/20^8^, followed by those over 55 on 12/01/21^9^, those over 35 on 28/01/21^10^ and everyone over the age of 16 since 03/02/2021^11^. Until early March 2020, individuals who had evidence of COVID-19 infection and had recovered were not eligible for vaccination. In early March 2020, they became eligible for a single dose. In addition to age eligibility, emergency, healthcare, defense forces and education workers and in late January also high school pupils have been prioritized for vaccination^8,12^. Finally, resourceful individuals have at times succeeded in getting vaccinated outside of eligibility criteria, for example by waiting for leftover doses outside of clinics. By 04/01/21, for example, more than 30% of vaccinated individuals were under the age of 60^13^ despite not being formally eligible.

The three largest population groups in Israel are the non ultra-orthodox Jewish population (thereafter called “general Jewish”), and the Ultra-Orthodox Jewish and Arab (mainly Muslim and Christian) populations. These groups comprise the Israeli population alongside smaller groups such as the Druze and Circassians. Each community tends to live in separate localities, and compared with the general Jewish population, Ultra-Orthodox and Arab populations are socio-economically deprived^14^. In addition these two populations are much younger than the general Jewish population: in 2015 29% of the general Jewish population was under 18, compared with 56% of Ultra-Orthodox Jews and 43% of Arabs^15^ (table 1).

**Table 1:**
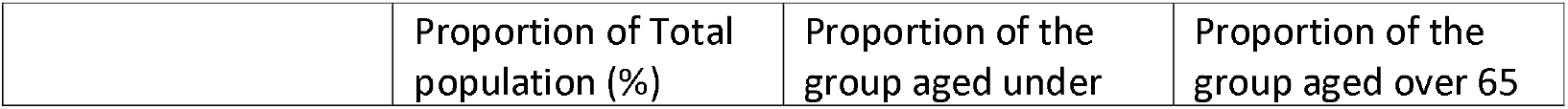
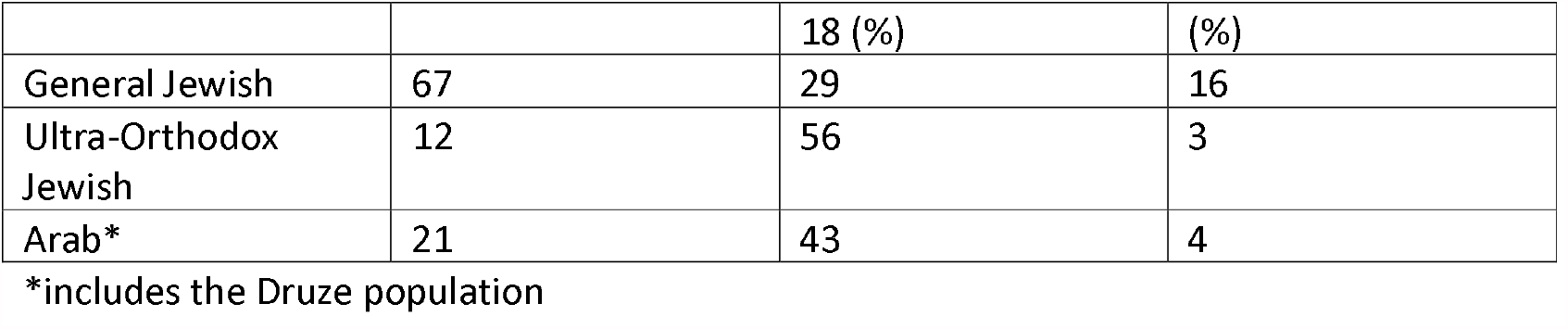
Age Distribution by population group, Israel, 2015^15^.

|  | Proportion of Total population (%) | Proportion of the group aged under | Proportion of the group aged over 65 |
| --- | --- | --- | --- |

|  |  | 18 (%) | (%) |
| --- | --- | --- | --- |
| General Jewish | 67 | 29 | 16 |
| Ultra-Orthodox Jewish | 12 | 56 | 3 |
| Arab* | 21 | 43 | 4 |

\*includes the Druze population

Despite the rapid pace of vaccination in Israel, vaccination coverage varies widely by geography, and therefore population group. By April 10 2021, vaccination coverage by municipality for the first dose ranged 1.97-90.12%^6^. These disparities exist despite local availability of the vaccine in the entire country and free vaccination for all. Comparing crude coverage between localities and population groups may however be misleading because of the very different age structure underlying each group and because COVID19 incidence varied by locality. In order to describe inequalities in COVID19 vaccine initiation we determined differences in age-specific dose 1 vaccine coverage between the main population groups in Israel, and characterised the influence of age on vaccine coverage within each of these groups.

## Methods

We generated a dataset of dose-1 vaccine coverage by age group and by municipality, by linking two publicly available datasets: first, population size by municipality in 5 year age-bands^16^; The second is the daily cumulative number of first doses of COVID19 given in each municipality, by age group ^17^. The datasets were linked using municipality codes unique to each locality. We categorized the population in 6 age groups: 15-19, 20-29, 30-39, 40-49, 50-59 and 60 and over. The most recent available population census with age distribution was from 2018^18^. In order to adjust for population growth, we extrapolated the population in each age group by the growth factor for 2018-2021.

Sectors were categorized using the municipality form code in the population size dataset to determine settlements with an Arab (including Druze) majority. There is no official category identifying Ultra-orthodox municipalities, so we set an arbitrary criterion whereby municipalities where Ultra-orthodox residents comprised 60% or more of the population was categorized as Ultra-orthodox. Municipalities with an Ultra-Orthodox population between 40 and 59% were considered mixed Jewish.

A 4 (sector: Arab, General Jewish, Ultra-Orthodox Jewish and Mixed Jewish) by 6 (age group: 15-19, 20-29, 30-39, 40-49, 50-59, 60+) two-way ANOVA test, was performed for the differences between sectors, age groups and interaction in order to determine whether the relative proportion of people vaccinated in each age group changed by population group. The analysis was weighted so that each age group in each municipality was adjusted to represent its relative proportion within each population group or age group.

In order to quantify the relative differences in vaccine coverage between population groups within each age group, and between age groups within each population, we conducted a series of binomial regressions. The “Mixed Jewish” category was excluded from the regression analysis because of the challenges in interpreting the output, as population in these municipalities is too heterogeneous to provide useful information about specific population groups

Analyses were conducted using R and STATA16 (Sata corp, Texas). The analysis made exclusive use of publicly available data and no ethics approval was required.

## Results

A total of 8,507,723 individuals in 268 cities, towns and villages with valid information were included. Of these 97 municipalities were Arab (including Druze) representing 16.6% of the sample population (table 2), 11 were Ultra-Orthodox (7.1% of the sample population), 154 as General Jewish (63.2%) and 6 as mixed (13.1%), including Jerusalem, Israel’s largest city.

**Table 2.** Differences in COVID 19 vaccination by population group among each age group, Israel, April 2021. All risk ratios are statistically significant, p<0.001

| Age group | Population group | Number eligible (n) | Vaccine coverage (%) | RR (within age groups) | 95%CI |
| --- | --- | --- | --- | --- | --- |
| 60+ | General Jewish | 1021937 | **93.4** | baseline |  |
|  | Ultra-Orthodox Jewish | 32451 | **81** | 0.867 | 0.863-0.872 |
|  | Arab | 107170 | **86.6** | 0.928 | 0.925-0.930 |
| 50-59 | General Jewish | 519195 | **87.8** | baseline |  |
|  | Ultra-Orthodox Jewish | 23831 | **64.7** | 0.737 | 0.730-0.744 |
|  | Arab | 114842 | **79.1** | 0.902 | 0.899-0.905 |
| 40-49 | General Jewish | 645759 | **81.7** | baseline |  |
|  | Ultra-Orthodox Jewish | 42039 | **55.5** | 0.679 | 0.673-0.685 |
|  | Arab | 153143 | **72.3** | 0.885 | 0.882-0.887 |
| 30-39 | General Jewish | 659082 | **76.4** | baseline |  |
|  | Ultra-Orthodox Jewish | 65256 | **48.8** | 0.639 | 0.634-0.644 |
|  | Arab | 165208 | **71.2** | 0.932 | 0.929-0.935 |
| 20-29 | General Jewish | 642710 | **77.8** | baseline |  |
|  | Ultra-Orthodox Jewish | 82097 | **48.1** | 0.618 | 0.614-0.623 |
|  | Arab | 237135 | **67.9** | 0.872 | 0.870-0.875 |
| 15-19 | General Jewish | 362325 | **50.4** | baseline |  |
|  | Ultra-Orthodox Jewish | 58667 | **26.6** | 0.528 | 0.521-0.536 |
|  | Arab | 142887 | **38.5** | 0.764 | 0.759-0.770 |

Vaccine coverage decreased with age (table 2, figure 1) and the difference between age groups was significant within all population groups ((F(5,1584) = 1137.96, p < 0.001, η^2^_p_ = 0.78)). Across all age groups, coverage was lower in other population groups compared with the general Jewish population (Figure 1, Table 2) and the differences were statistically significant ((F(3,1584) = 626.75, p < 0.001, η^2^_p_ = 0.54). Gaps between population groups differed according to age group (F(15,1584) = 11.49, p < 0.001, η^2^_p_ = 0.10).

**Figure 1:**
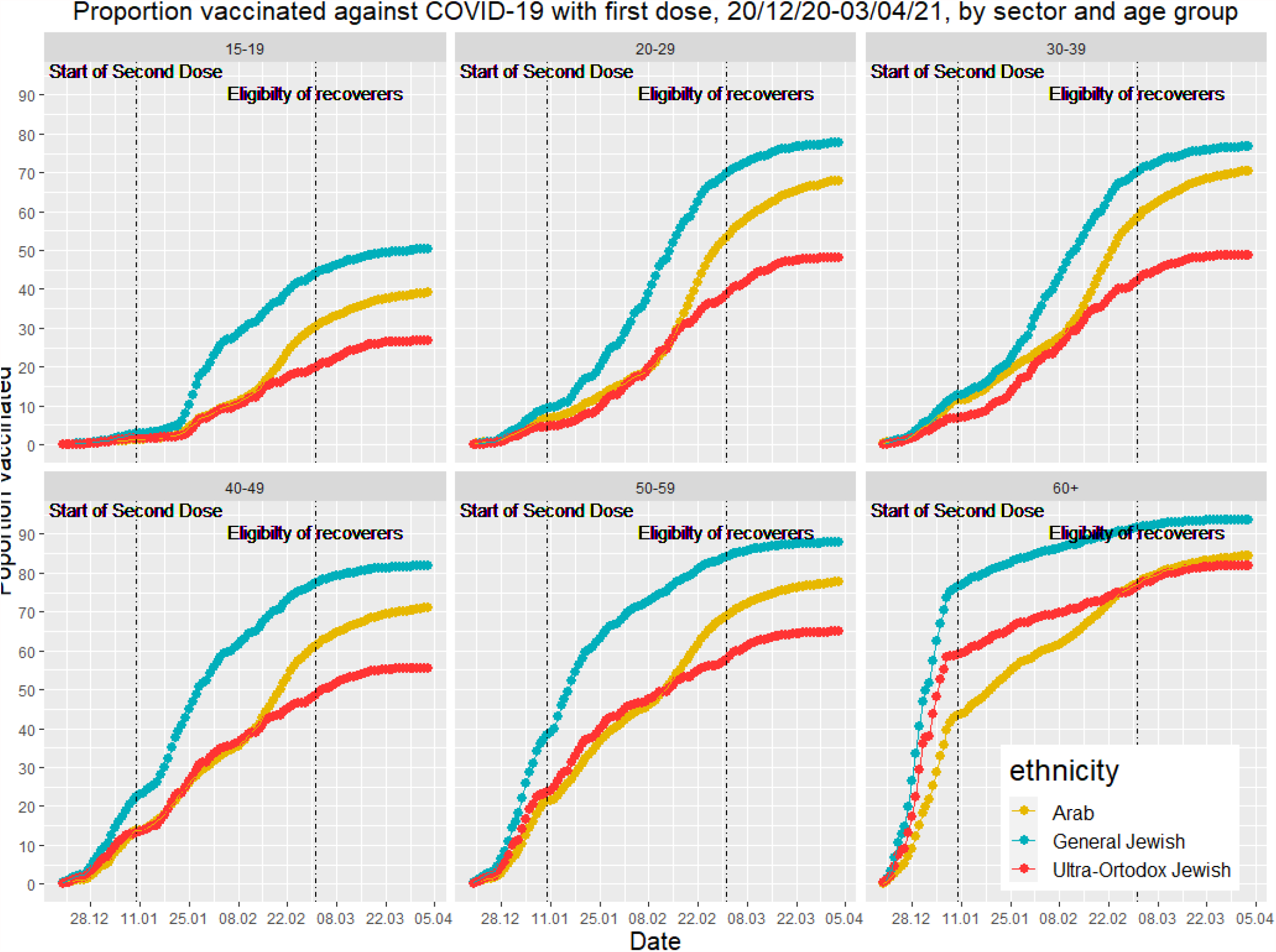
First dose coverage of COVID-19 vaccination by age group and sector in Israel, 20/12/20 - 03/04/21

Compared with coverage the general Jewish population, coverage was lowest in the Ultra-Orthodox population in all age groups (range -13.3%, 95%CI -13.7%;-12.8% among 60+ to - 47.2% 95%CI -47.9%; -46.4% among 15-19 years old, p<0.001, table 2). Within each population group, the proportions of individuals in younger age groups vaccinated relative to older age groups differed. They decreased with decreasing age in all groups and were smaller in the Ultra-Orthodox groups compared with the General Jewish and Arab populations (figure 2, table 3). For example within the general Jewish population, people aged 20-29 were 16.6% less likely to be vaccinated than those over the age of 60, increasing to 40.6% in the Ultra Orthodox population (figure 2, table 3). The Ultra-orthodox population was the least vaccinated in all age groups relatively to those aged 60 and over (figure 2)

**Figure 2:**
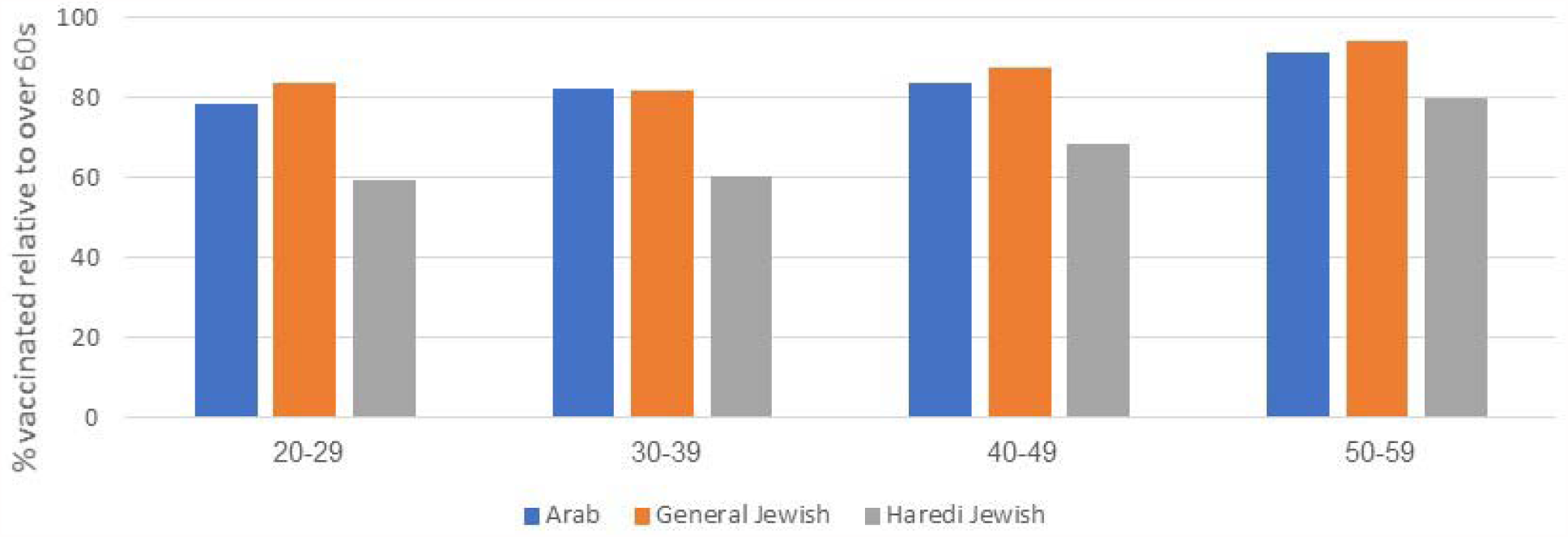
Proportion vaccinated against COVID19 relative to those over the age 60, by age group, Isreal, April 2020

**Table 3:**
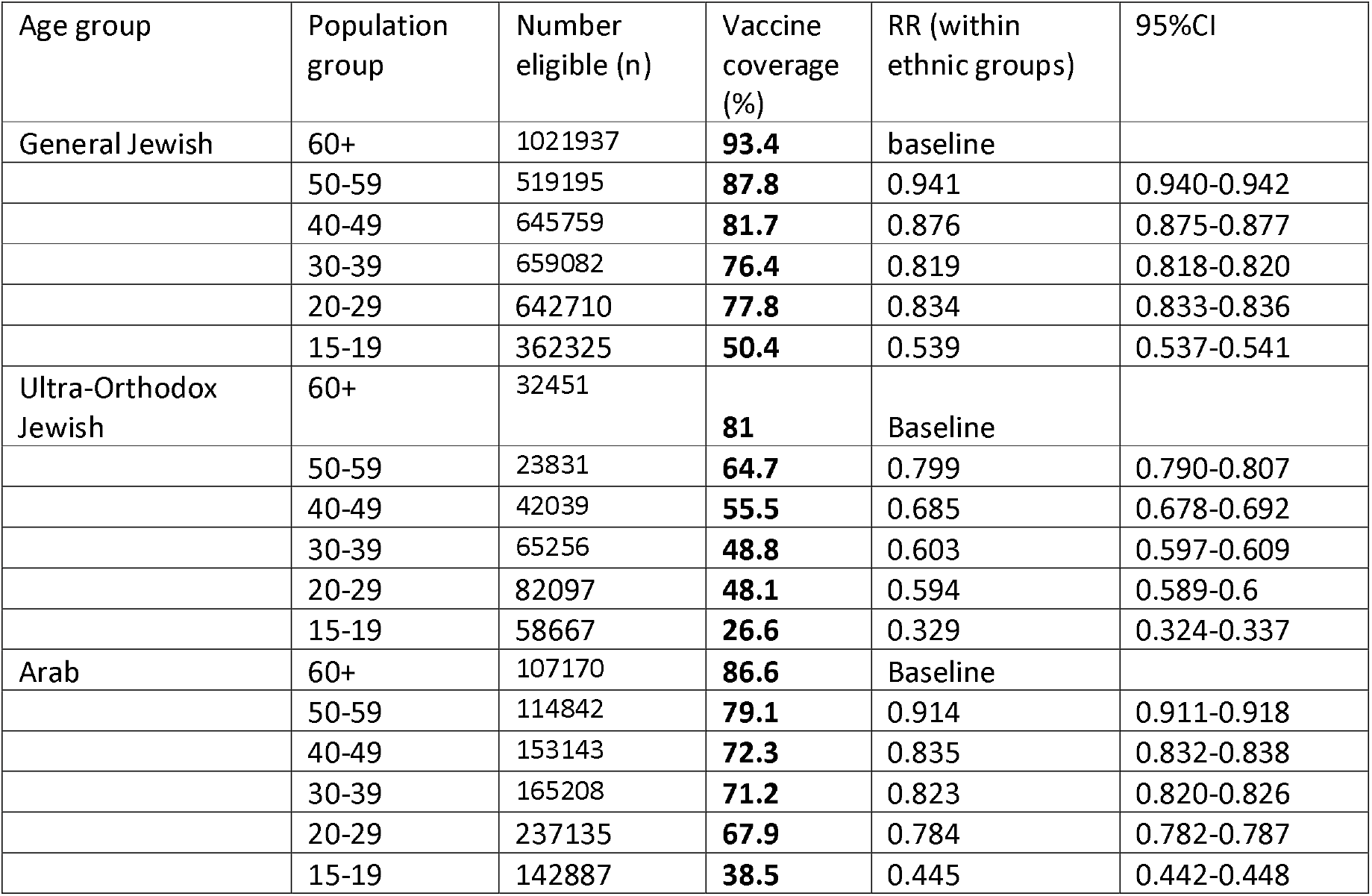
Differences in COVID 19 vaccination by age group among each population group, Israel, April 2021. All risk ratios are statistically significant, p<0.001

## Discussion

Coverage for the COVID19 vaccine in Israel, varies widely in different groups comprising Israeli society. In all age groups, the Ultra-Orthodox population had the lowest vaccine coverage. Vaccine coverage was low among individuals aged 15-19, and younger individuals in the Ultra-Orthodox population were the least likely to be vaccinated, with the size of the relative differences between this group and others increasing with decreasing age, and the relative difference in vaccine coverage between younger and older age groups being larger in the Ultra -Orthodox population than in the general Jewish or Arab population. Expressed differently, The younger the age group, the more Ultra-Orthodox Jews are diverging from their age peers in terms of initiating COVID19 vaccination. These findings suggest generational differences in terms of vaccination behaviour in this group. This study does not provide an explanation for this observation, but identifying age and population group specific barriers to vaccination, and providing tailored strategies to increase vaccination in this group is an urgent priority considering the uncertainty over the pandemic. Low vaccination in this group could initially be explained by high incidence of COVID19 among young Ultra-Orthodox Jews because infected individuals were not eligible. However they became eligible in March 2021. Incidence was also high among young individuals in the Arab population, yet coverage in this group increased and became much closer to similar age groups in the general Jewish population compared with the Ultra-Orthodox groups. Tailored approaches to vaccination are widely recognized as an effective approach to successful vaccination programmes in minority groups.

Our study demonstrates how coverage figures can be misinterpreted if interpreted at face value because of the very different age structures and background incidence in the different populations included in this study. While an age-stratified analysis shows that among different ethno-religious groups differences in those over the age of are relatively small, other groups especially younger Ultra-Orthodox groups remain under-vaccinated. This is a threat to achieving full herd immunity against COVID19 through vaccination, which will require equitable coverage in all population groups, in particular in a small, densely populated country such as Israel. If the Ultra-Orthodox population under 40 remains undervaccinated, the risk of recurrent clusters of COVID19 in this population is high, in particular as international travel reopens and high volume of travel between Ultra-Orthodox communities in Israel and abroad resumes. This epidemiological picture has been described in the Ultra-Orthodox communities for other infectious diseases such as measles^19^. As vaccine eligibility extends to children, as is planned in Israel, the impact of an undervaccinated Ultra-Orthodox population may be compounded by the fact that the proportion of children under the age of 18 is higher in this group than any other population group in Israel, and most parents will be under the age of 40, the least vaccinated group. Although our study does not measure parent’s intention to vaccinate their children, evidence suggests that parent’s intention to vaccinate their children against COVID is lower than their intention to vaccinate themselves.^20^ While it is important to identify and address immunity gaps in the most underserved groups, continued efforts to improve coverage is essential, in particular in younger individuals where coverage remains low in all populations groups.

### Strengths, limitations and further directions

This study is to our knowledge the first to describe how age and belonging to a particular group interact to influence COVID19 vaccine coverage in Israel. Our study has used comprehensive datasets taken from official governmental databases, covering the majority of the population is Israel. Bias and representativeness are therefore unlikely to be issues. We excluded municipalities in which the population was too diverse to assign it to a particular category. This concerns a large proportion of the population, including Jerusalem, Israel’s largest city.

There are several limitations to this study. First, the most recent demographic data was from 2018, which leads to a slight underestimation of the denominator and over estimation of vaccine coverage. However the timeframe between 2017 and 2021 is too short to significantly impact on the relative distribution of the population by age or population group, and relative measures are therefore unlikely to be significantly affected. We applied a correction factor based on national growth to try and minimize the effect of an underestimated denominator. However a few cities have undergone extensive growth since 2017. This made the denominator (i.e., population in 2017) smaller than the numerator (i.e., number of people vaccinated) in those cities. Data for these municipalities was uninterpretable and therefore excluded. We also made the assumption that the population groups considered in the study (General Jewish, Ultra-Orthodox and Arab) are homogenous in the vaccination behaviour, which is unlikely to be true. More localized analyses, combined with qualitative research understanding barriers and enablers in specific communities, will enable to understand drivers of vaccination with more granularity.

Another limitation was that the exact numbers of vaccination administered was suppressed when it was less then 15. As a pragmatic measure, we assigned 10 vaccines when less than 15 was indicated. Although not exact, the small numbers compared with the overall large sample size are unlikely to bias the findings.

Another limitation is that the proportion of Ultra-Orthodox population (7.1%) and Arab population (16.6%) is smaller in the study sample than in the population (12% and 21% respectively). This is mostly because significant proportions of these populations live in municipalities counted as General Jewish or mixed (30% of ultra-Orthodox and 10% of Arab population); and because of the exclusion of Jerusalem, home to over 24% of the Ultra-Orthodox population 19% of the Arab population

Our analysis offers an in depth characterization of how age, population groups and their interplay impact on vaccine coverage. As of April 11^th^ 2021 Israel remains the country with the highest proportion of its population who has initiated vaccination against COVID19 with a first dose. Nevertheless the rate of coverage increase is slowing down as those eager to be vaccinated have now received the vaccine. The vaccination progamme now requires tailored approaches in order to convince those who are more indifferent or perhaps even skeptical about vaccination^20^, to get vaccinated. Our study identifies a number of groups, in particular younger individuals in general but also specifically in the Ultra-orthodox population, as falling behind others in terms of vaccine coverage. Qualitative studies understanding the causes behind this divergence are urgently needed to inform tailored vaccination strategies.

## Data Availability

All data is publicly available at https://data.gov.il/dataset/covid-19

